# Hepatic resection versus transarterial chemoembolization for the intermediate stage hepatocellular carcinoma: A cohort study

**DOI:** 10.1101/2020.10.18.20214833

**Authors:** Linbin Lu, Peichan Zheng, Zhixian Wu, Xiong Chen

## Abstract

**Background:** The selection criterion for hepatic resection(HR) in intermediate-stage(IM) hepatocellular carcinoma(HCC) is still controversial. We used real-world data to evaluate the overall survival (OS) treated with HR or TACE.

**Methods:** In all, 942 patients with IM-HCC were categorized in HR and TACE groups. OS was analyzed using the Kaplan-Meier method, log-rank test, Cox proportional hazards models, and propensity score-matched (PSM) analyses. The smooth curve was performed through the generalized additive model. The interaction test was performed to evaluate the HR impact on OS concerning risk factors. Also, we used multiple imputation to deal with the missing data.

**Results:** Totally, 23.0% (n=225) of patients received HR. At a median overall survival of 23.7 months, HR was associated with the improved OS on multivariate analysis (hazard ratio, 0.45; 95%CI: 0.35, 0.58; after PSM: 0.56; 95%CI: 0.41, 0.77). Landmark analyses limited to long-term survivors of ≥ 6 months, ≥ 1, and ≥ 2 years demonstrated better OS with HR in all subsets (all *P*<0.05). After PSM analysis, however, HR increased 20% risk of death (HR, 1.20; 95%CI: 0.67, 2.15) in the subgroup of LDH ≤192 U/L (*P* for interaction = 0.037). Furthermore, the significant interaction was robust between the LDH and the HR with respect to 1-, 3-, and 5-year observed survival rate (all P<0.05).

**Conclusion:** Hepatic resection was superior to TACE for intermediate-stage HCC in the range of LDH level > 192 U/L. Moreover, TACE might be suitable for patients with LDH level ≤ 192 U/L.

**Synopsis:**

- Hepatectomy was superior to TACE for BCLC-B HCC.
- Hepatectomy increased 20% risk of death for LDH < 192 U/L after matching.
- A significant interaction was robust between LDH and with respect to hepatectomy the 1-, 3-, and 5-year observed survival rate.

## 1. Introduction

Hepatocellular carcinoma (HCC) is one of the most leading causes of cancer-related death worldwide and the fifth cause of death in China[1]. According to the BCLC staging system, the most widely used scheme, patients with early stages (stage 0 and A) are suitable for hepatic resection (HR), while intermediate-stage (IM) HCC patients are recommended for transarterial chemoembolization (TACE)[2]. Compared with conservative treatment for IM-stage (stage B) HCC, patients treated with TACE have a better 2-year overall survival[3]. After selecting the Bolondi’s criteria[4], the patients with stage B1 or B2 have a higher 5-year survival rate (21.4% vs. 13.9%)[5]. Subsequently, the subgroup of IM-stage HCC patients who benefit from TACE was identified through numerous criteria, including the ART score[6], ABCR score[7], the ALBI grade[8], etc. Although the highly selected HCC patients have a median survival of 51.5 months[9], TACE’s role is challenged by hepatic resection(HR).

A meta-analysis including 18 high-quality studies is recently performed to compared survival outcomes of 5,986 patients after HR and TACE. They find that both stage B and stage C patients show significantly better overall survival for HR than TACE[10]. However, the controversial evidence has emerged that HR is superior to TACE only in the lower mortality risk subgroup of IM-stage HCC [11; 12; 13; 14; 15], such as BCLC stage B1/B2[12; 13]. Although the subgroup of IM-stage HCC has been selected by the predicted models with a median overall survival of 61.3 months, patients who are more suitable for HR are still controversial. Interestingly, Cucchetti et al. [16] perform a regret-based decision curve analysis (Regret-DCA) to choose HR or TACE for IM-stage HCC. In this study, HR should be offered to the patients with 3yr mortality risk<35%, but the optimal strategy (HR vs. TACE) is still unclear when the mortality risk between 35% and 70%. Although numerous subgroups have been identified, more promising biomarkers are urgent to choose better therapy.

To deal with this issue, we construct a real-world propensity score-matched cohort study to compare hepatic resection and transarterial chemoembolization in the intermediate-stage hepatocellular carcinoma,

## 2. Methods and patients

### 2.1 Patient selection

Clinical and biological data in our study had been previously published in full[17]. In this study, we mainly focus on the derivation cohort from Sun Yat-sen University Cancer Center(SYSUCC) between January 2007 and May 2012. The details of inclusion criteria were shown in Fig S1. A total of 979 patients were included in the derivation cohort. In the derivation cohort, 37(37/979, 3.8%) patients were excluded for refusing to receive treatment, and 942 patients were included into final analysis, with TACE (717/979, 73.2%) or surgical resection (225/979, 23.0%) as their first-line treatment. A total of 805 patients were afforded after initial treatment at the second follow-up visit (n=597 after TACE, n= 208 after HR). According to the decision form multidisciplinary teams, the second-line therapy include ablative therapies (n=66/805, 8.2%), surgical resection (n=38/805, 4.7%), repeated TACE (n=172/805, 21.4%), other therapies (n=5/805, 0.6%), or best supportive care (n=524/805, 65.1%).

The Ethics Committee of SYSUCC approved the study protocol (2017-FXY-129). Because this was a retrospective study, the informed consent was waived.

### 2.2 Diagnosis, treatment and follow-up

For the patients treated with HR, HCC diagnosis was confirmed by histopathological examination of surgical samples. In contrast, for the patients with TACE, the diagnosis was established by the combination with the serum level of alpha-fetoprotein (AFP, over 400 ng/mL) and clinical imaging, including ultrasonography, computed tomography, or magnetic resonance imaging. If the diagnosis was uncertain based on imaging and AFP level, a needle biopsy was performed.

Based on the multidisciplinary teams’ decisions, the optimal treatment plans were adopted for each HCC patient. Indications for HR in the IM-HCC patients were appropriate residual liver volume determined by computed tomography. For the patients without cirrhosis, 30% remnant liver volume after HR was considered adequate. However, for those with chronic hepatitis, cirrhosis, and severe fatty liver, the remnant volume should be more than 50%. Liver resection should not be carried out among intermediate or advanced cirrhosis patients and poor liver function (Child-Pugh C). Patients who satisfied the indications for HR were treated by surgical resection unless the patient requested TACE.

During the initial treatment period for the first two years, patients were followed up for every 2 or 3 months if complete remission was achieved. The frequency gradually decreased to every 3 to 6 months after two years’ remission.

### 2.3 Variables and definition

Patients were stratified as the group of hepatic resection (HR) and transarterial chemoembolization (TACE). HR was defined as surgical therapy for the lesions in hepatic segments or lobes. In the clinical, patients with well liver function and less tumor loading were usually suitable for HR. TACE was defined as chemoembolization of the hepatic artery. Categorical variables consisted of gender, Child-Pugh class (A, B), intrahepatic tumor number(⩽3, >3), both lobe with lesions (no, yes). Continuous variables, including age, the diameter of the main tumor, alpha-fetoprotein (AFP), C-reactive protein (CRP), lactate dehydrogenase (LDH), hemoglobin (Hgb), white blood cell count (WBC), and platelet count (PLT) level, were also regarded as categorical variables. AFP and PLT were transformed into the Log_10_ scale because of their left skew. All variables were afforded at the baseline before any anti-cancer treatment. The endpoint of interest was overall survival (OS), which was defined as the time from diagnosis to death by any cause. BCLC stage B and CNLC (China liver cancer staging) HCC were defined as follows [18; 19]: BCLC Stage B: Two to three lesions, at least 1 is more than 3 cm in diameter; or more than 3 lesions of any diameter. ECOG PS 0. Child-Pugh class A or B. Without blood vessel invasion and extrahepatic metastases.

CNLC stage IIa: Two to three lesions, of which at least 1 is more than 3 cm in diameter. ECOG PS 0-2. Child-Pugh class A or B. Without Blood vessel invasion and extrahepatic metastases.

CNLC stage IIb: More than three lesions of any diameter. ECOG PS 0-2. Child-Pugh class A or B. Without Blood vessel invasion and extrahepatic metastases.

### 2.4 Statistical analyses

To compare the differences of baseline characteristics between the HR and TACE groups, we compared categorical variables with the chi-square test and continuous variables by the Mann-Whitney test.

Firstly, Survival was calculated using the Kaplan-Meier method, and univariate comparisons were performed using the log-rank test and unadjusted Cox models. Also, multivariable Cox proportional hazards models were adjusted for factors, including Child-Pugh class, the diameter of main tumor, location of lesions, intrahepatic tumor number, AFP, LDH, and PLT level.

Next, to account for potential biases favoring the administration of HR to patients with more favorable baseline prognoses, sequential landmark analyses were performed to evaluate survival with HR or TACE for patients with a minimum of ≥ 6 months, ≥ 1 year, and ≥ 2 years survival from diagnosis. Interaction and stratified analyses were performed for the covariates selected a priori, including Child-Pugh class, diameter of main tumor, location of Lesions, intrahepatic tumor number, CNLC stage, AFP, LDH, and PLT level. To further explore the interaction, the smooth curve was performed between Log LDH and observed mortality at 1, 3, and 5 years through the generalized additive model.

### 2.5 Sensitivity analysis

Finally, we conducted three approaches to evaluate the core result as a sensitivity analysis. To minimize the potential bias, propensity score (PS)-matched analyses were performed to compare TACE or HR outcomes. One-to-one matching (TACE vs. HR) without replacement was completed using the nearest neighbor match on the logit of the PS (derived from age, diameter of main tumor, location of lesions, intrahepatic tumor number, AFP, Hgb, LDH, WBC, CRP, all P<0.05 in Table 1). The caliper width was 0.02 times the standard deviation of the logit of the propensity score.

**Table 1.** Baseline characteristics between TACE and HR group in the derivation cohort.

|  | Treatment |  | P-value |
| --- | --- | --- | --- |
|  | TACE (n = 717) | HR (n = 225) |  |
| Age | 53.9 ± 12.3 | 50.9 ± 12.6 | 0.001 |
| Gender |  |  | 0.802 |
| Male | 654 (91.2%) | 204 (90.7%) |  |
| Female | 63 (8.8%) | 21 (9.3%) |  |
| HBV infection |  |  | 0.132 |
| No | 18 (2.8%) | 2 (1.0%) |  |
| Yes | 622 (97.2%) | 203 (99.0%) |  |
| Child-Pugh class |  |  | 0.302 |
| A | 613 (85.5%) | 186 (82.7%) |  |
| B | 104 (14.5%) | 39 (17.3%) |  |
| Diameter of main tumor(cm) | 7. 5± 3.8 | 6.4 ± 2.8 | <0.001 |
| Location of lesions |  |  | <0.001 |
| Unilobar | 254 (35.4%) | 148 (65.8%) |  |
| Bilobar | 463 (64.6%) | 77 (34.2%) |  |
| Intrahepatic tumor number |  |  | <0.001 |
| ≤3 | 237 (33.1%) | 142 (63.1%) |  |
| >3 | 480 (66.9%) | 83 (36.9%) |  |
| AFP(ng/ml) |  |  | 0.014 |
| <25 | 180 (26.5%) | 75 (35.2%) |  |
| ≥25 | 500 (73.5%) | 138 (64.8%) |  |
| CRP(mg/L) |  |  | <0.001 |
| <10 | 318 (45.4%) | 72 (32.4%) |  |
| ≥10 | 382 (54.6%) | 150 (67.6%) |  |
| Hgb(g/L) |  |  | <0.001 |
| <120 | 148 (20.8%) | 71 (31.6%) |  |
| ≥120 | 562 (79.2%) | 154 (68.4%) |  |
| LDH(U/L) |  |  | <0.001 |
| <245 | 356 (50.1%) | 152 (67.6%) |  |
| ≥245 | 354 (49.9%) | 73 (32.4%) |  |
| WBC(10<sup>9</sup>/L) |  |  | <0.001 |
| <11 | 611 (86.9%) | 162 (74.0%) |  |
| ≥11 | 92 (13.1%) | 57 (26.0%) |  |
| PLT(10<sup>9</sup>/L) |  |  | 0.249 |
| <150 | 381 (53.7%) | 111 (49.3%) |  |
| ≥150 | 328 (46.3%) | 114 (50.7%) |  |

Numbers that do not add up to 942 are attributable to missing data. The chi-square test was performed for categorical measures and Kruskal-Wallis Test for continuous measures . \*Fisher's exact probability test.

We also used multiple imputation (MI) to maximize statistical power and eliminate bias, which may occur if the confounders with missing data were excluded from the analysis. The MI was based on five replications and the Markov-chain Monte Carlo method in the R MI procedure, to account for missing data on Child-Pugh class, diameter of main tumor, location of lesions, intrahepatic tumor number, PLT, Child-Pugh class, AFP, and LDH. We then conducted an MI cohort to performed sensitivity analyses using a complete-case analysis.

To eliminate the effect of ablative therapies and surgical resection in the second-line treatment, we build a secondary cohort based on the MI cohort without those therapies. All the multivariable Cox analyses mentioned above were repeated in the PS, MI, and secondary cohort.

Statistical analysis was performed using Empower (www.empowerstats.com, X&Y solutions, inc. Boston MA) and R software (version 3.4.3). P-value < 0.05 considered significant.

## 3. Results

### 3.1 Descriptive characteristics

After excluding those who refused to receive treatment(n=33), a total of 942 HCC patients were included in the derivation cohort, including 563 patients (59.8%) with CNLC stage IIb (480 patients for TACE, 83 patients for HR), 379 patients (40.2%) with stage IIa (237 patients for TACE, 142 patients for HR). All patients had well performance status (ECOG PS 0). After first-line treatment with TACE, 46 patients(6.6%, 46/597) had the invasion of portal vein or its branch (n=38), hepatic veins(n=6) and Vena Canva/Atrium(n=2), 53 patients(8.9%) with distant metastasis, while 36 patients(6.0%) with lymph node metastasis at the second follow-up visit.

In the derivation cohort, patients with HR were younger, the shorter diameter of the main tumor, lower hematological indicators (AFP, CRP, Hgb, LDH, WBC), less frequently intrahepatic tumor number, and both lobe with lesions(all p<0.05), which was shown in Table 1. The majority of the patient (825/942,87.6%) had hepatitis B virus (HBV) infection, who was treated with nucleos(t)ide analog therapy. The difference in HBV infection rate was not significant between the HR and TACE group.

### 3.2 Survival analysis for the entire cohort

As shown in Fig 1, the median overall survival (mOS) for the entire cohort was 23.7 (95%CI: 20.4, 27.2) months. The mOS is 18.5 (95%CI:16.9, 20.3) months for the TACE group versus 67.4 (95%CI: 46.7, NA) months for the HR group (*P*<0.0001). After PS-matching, the difference of mOS between TACE (29.9 months, 95%CI: 22.5, 38.9) and HR group (67.4 months, 95%CI: 44, NA) is still significant (*P*<0.0003).

**Fig 1.** Kaplan-Meier curves of overall survival in the derivation cohort stratified with HR and TACE. (A) All patients. (B) Propensity score-matched patients.

In univariable analysis focusing on the entire cohort (Table 2), Child-Pugh class(vs A: HR, 1.28; 95%CI: 1.01, 1.62), diameter of main tumor(vs <5: HR, 2.28; 95%CI: 1.86, 2.80), location of lesions(vs unilobar: HR, 1.50; 95%CI:1.26, 1.79), intrahepatic tumor number(vs ≤3: HR, 1.55; 95%CI: 1.30, 1.86), AFP(vs <25: HR, 1.63; 95%CI: 1.33, 2.00), LDH (vs <245; HR,1.61; 95%CI: 1.36, 1.92) and PLT(vs <150, HR, 1.33; 95%CI: 1.12, 1.57) were significantly associcated with survival(all *P* <0.05). These variables were taken into further analysis.

**Table 2.**
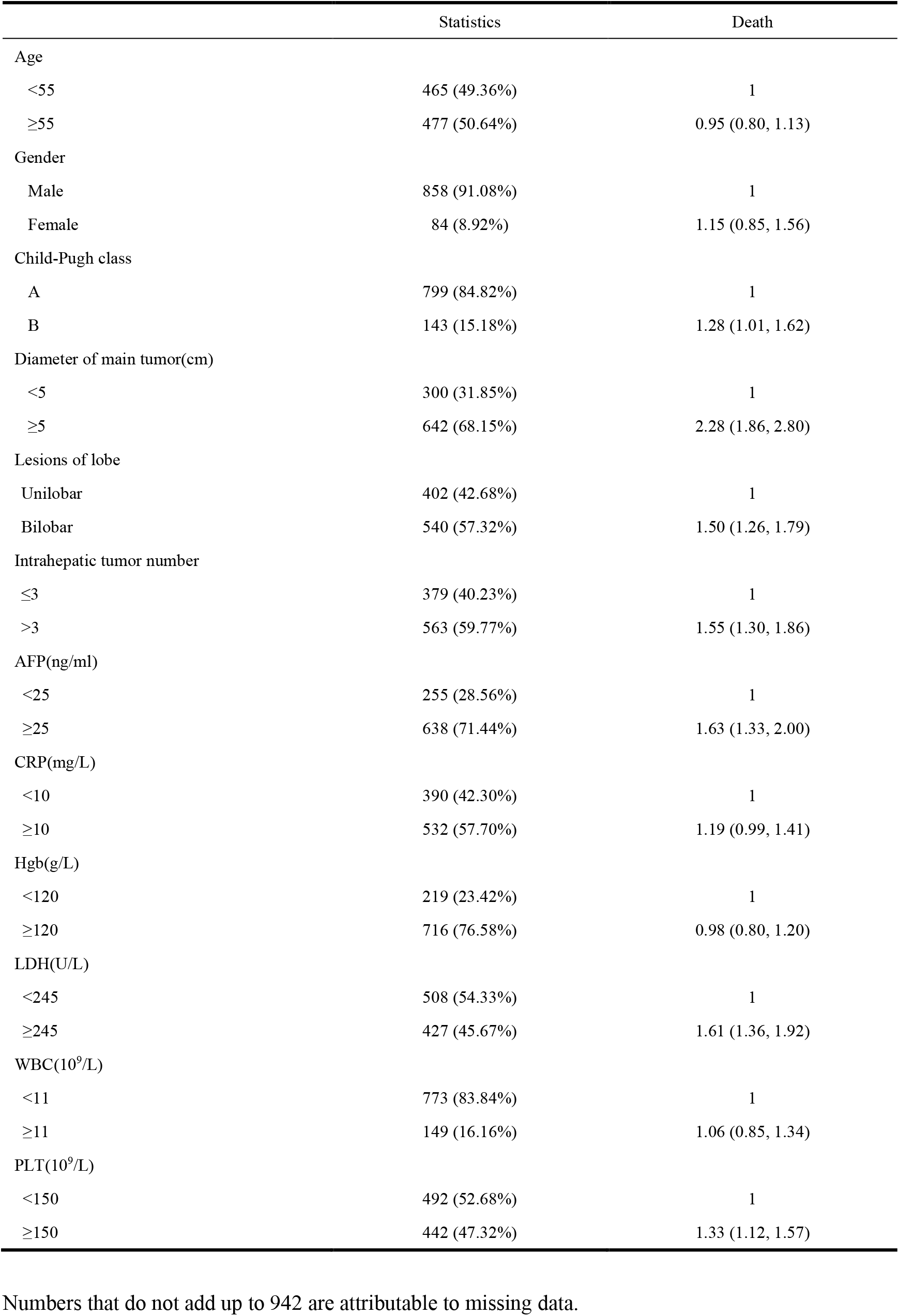
Univariate analysis of prognostic factors in the derivation cohort.

|  | Statistics | Death |
| --- | --- | --- |
| Age |  |  |
| <55 | 465 (49.36%) | 1 |
| ≥55 | 477 (50.64%) | 0.95 (0.80, 1.13) |
| Gender |  |  |
| Male | 858 (91.08%) | 1 |
| Female | 84 (8.92%) | 1.15 (0.85, 1.56) |
| Child-Pugh class |  |  |
| A | 799 (84.82%) | 1 |
| B | 143 (15.18%) | 1.28 (1.01, 1.62) |
| Diameter of main tumor(cm) |  |  |
| <5 | 300 (31.85%) | 1 |
| ≥5 | 642 (68.15%) | 2.28 (1.86, 2.80) |
| Lesions of lobe |  |  |
| Unilobar | 402 (42.68%) | 1 |
| Bilobar | 540 (57.32%) | 1.50 (1.26, 1.79) |
| Intrahepatic tumor number |  |  |
| ≤3 | 379 (40.23%) | 1 |
| >3 | 563 (59.77%) | 1.55 (1.30, 1.86) |
| AFP(ng/ml) |  |  |
| <25 | 255 (28.56%) | 1 |
| ≥25 | 638 (71.44%) | 1.63 (1.33, 2.00) |
| CRP(mg/L) |  |  |
| <10 | 390 (42.30%) | 1 |
| ≥10 | 532 (57.70%) | 1.19 (0.99, 1.41) |
| Hgb(g/L) |  |  |
| <120 | 219 (23.42%) | 1 |
| ≥120 | 716 (76.58%) | 0.98 (0.80, 1.20) |
| LDH(U/L) |  |  |
| <245 | 508 (54.33%) | 1 |
| ≥245 | 427 (45.67%) | 1.61 (1.36, 1.92) |
| WBC(10<sup>9</sup>/L) |  |  |
| <11 | 773 (83.84%) | 1 |
| ≥11 | 149 (16.16%) | 1.06 (0.85, 1.34) |
| PLT(10<sup>9</sup>/L) |  |  |
| <150 | 492 (52.68%) | 1 |
| ≥150 | 442 (47.32%) | 1.33 (1.12, 1.57) |

Numbers that do not add up to 942 are attributable to missing data.

Next, all of 7 variables were included in the multivariable analysis in Table 3. In Model I, adjusted HR (aHR) was 0.43 (95%CI: 0.34, 0.55) for liver resection compared to TACE. To explore the confounding factor’s nonlinearity, diameter of main tumor, log AFP, LDH, and log PLT were regarded as continuous variables in Model II. Compared with TACE, hepatectomy reduce 55% death risk (0.45; 95%CI: 0.35, 0.58). After PS-matching, hepatic resection was still superior to TACE (aHR, 0.56; 95%CI: 0.41, 0.77).

**Table 3.**
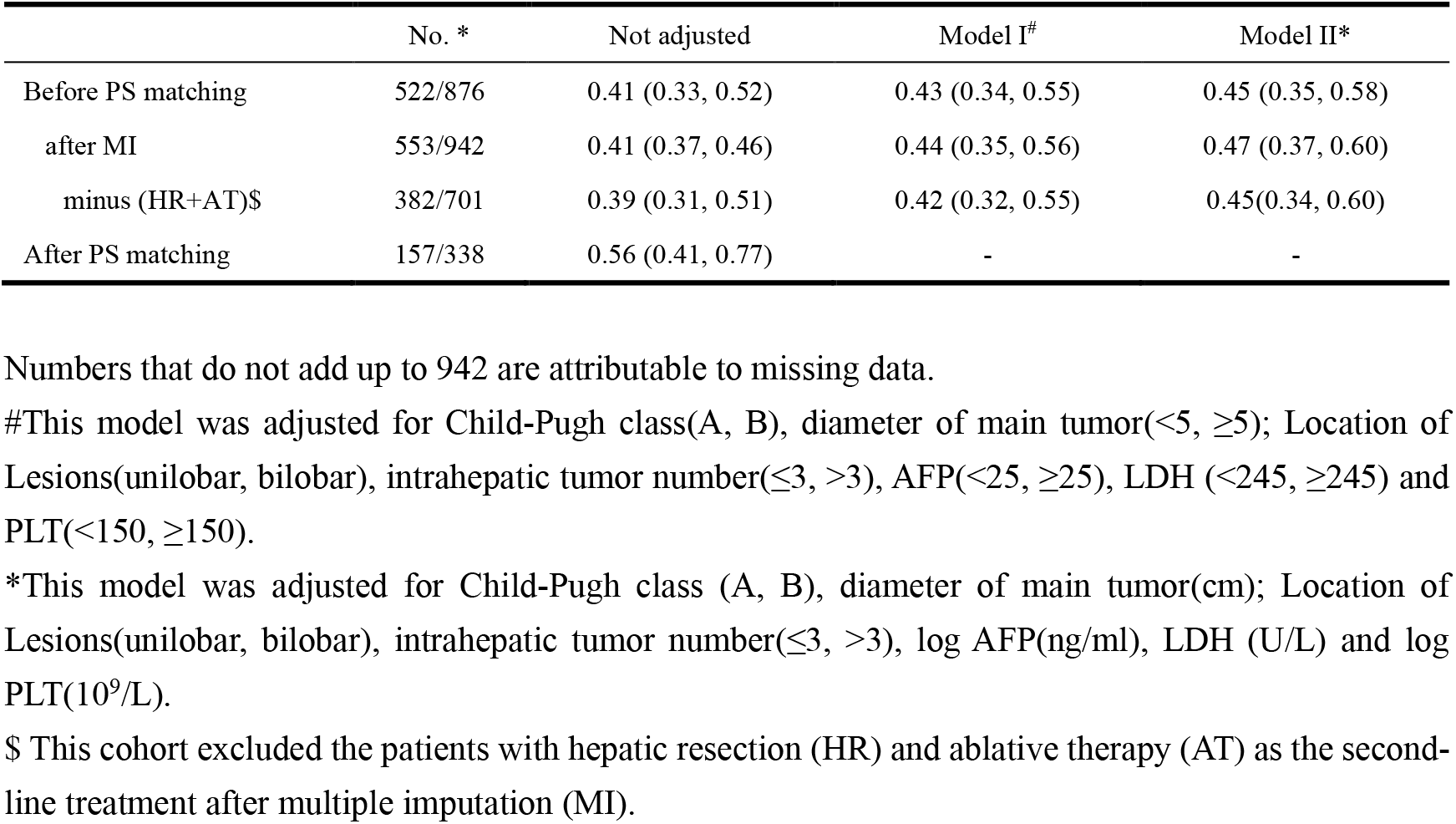
Hepatic resection (vs. TACE) and multivariate HRs of overall survival with 95% CIs in HCC with BCLC stage B.

Sequential landmark analysis revealed statistically significant improvement in OS with HR for patients surviving over 6 months (HR, 0.45; 95% CI: 0.35, 0.58), 1 year (HR, 0.46; 95% CI: 0.34, 0.62), and 2 year (HR, 0.52; 95%CI: 0.33, 0.79) (Fig 2, Table S2). On stratified analyses (Fig 3; Table S3-4), the magnitude of the association between HR and better survival was more significant for patients with higher LDH level (vs. bottom tertile; *P* for interaction = 0.006) and higher PLT (vs. bottom tertile; *P* for interaction = 0.037). After PS-matching, however, only for patients with higher LDH levels had significant interaction. In the subgroup of LDH < 192 U/L (bottom tertile), HR increased 20% risk of death (HR, 1.20; 95%CI: 0.67, 2.15). And HRs were 0.50 (95%CI: 0.30, 0.84) and 0.26 (95%CI: 0.14, 0.47) in the subgroup of middle tertile (192<LDH<255) and top tertile (LDH ≥ 255), respectively. No significant interactions were observed between the effect of TACE and Child-Pugh class, diameter of main tumor, location of Lesions, intrahepatic tumor number, CNLC stage, and AFP.

**Fig 2.** Landmark analyses of overall survival for long-term (≥ 6 months, ≥ 1 year, ≥ 2 years) survivors.

**Fig 3.** Association between overall survival and platelet count /lactic dehydrogenase stratified by tertile before(A)/after(B) PS-matching. (vs. TACE in the bottom tertile).

### 3.4 Sensitivity analysis

After multiple imputation, HR remained associated with the better OS using multivariable Cox regression on the imputed data set (Table 3). Adjusted HR was 0.44 (95%CI:0.35, 0.56) for Model I and 0.47 (95% CI: 0.37, 0.60) for Model II. Furthermore, the cohort results were still consistent in the MI cohort after excluding the patients with liver resection and ablative therapy as second-line treatment (Table 3).

After the PS-matching of the derivation cohort data set, there were no significant differences between the HR and TACE groups (both group, n = 169), shown in Table S1 and Fig S2. Median survival in the hepatic resection patients was 67.4 (95%CI: 44, NA) months and 29.9 (95% CI: 22.5, 38.9) months in TACE patients. Compared with TACE, liver resection continued to be associated with improved overall survival (HR, 0.56; 95%CI: 0.41, 0.77; P < 0.0003, Fig 1B). The C-statistic of the ROC–calculated PS was 0.66 (95%CI, 0.60, 0.72).

Besides, the 1-, 3-, and 5-year observed survival rate were 76.9%, 52.7%, 46.7% for TACE group and 85.8%, 68.6%, 63.3% for HR group. When the LDH level was less than 192 U/L, however, the mortality for HR patients was 2.89 (95%CI:0.71, 11.81) times, 1.20 (0.54, 2.65) times, and 1.22 (0.57, 2.62) times versus the TACE group at 1, 3, and 5 years (Table S4). The significant interaction was robust between the LDH and the HR concerning 1-, 3-, and 5-year observed survival rate (all *P* < 0.05, see Fig 4 and Table S5).

**Fig 4.** The smooth curve between Log LDH and observed mortality at 1, 3, and 5 years stratified with HR and TACE.

## 4. Discussion

In this large-scale, real-world data, we found that the overall survival for HR was significantly better than their TACE counterparts, which was consistent with the previous literature [10; 20]. Interestingly, Toshifumi et al. [11] also reported that liver resection reduced 44% death risk after PS matching (HR, 0.56). Notably, this finding remained marked after adjusting for crucial clinical confounders. When the LDH level increased, the magnitude of the association between liver resection and better survival was more significant. After PS matching, however, hepatic resection was associated with worse survival compared with TACE but not significantly. To the best of our knowledge, we first observed the significant interaction between the effect of HR and LDH levels.

TACE had been recommended as the first-line treatment for unresectable intermediate-stage HCC [19]. However, it remained a great controversy about whether surgery was recommended for the resectable BCLC-B HCC patients with good liver functional reserve. In the clinical practice from Asia–Pacific region[21], No. intrahepatic lesson more than three tumors, both lobes with tumors or satellite nodules were not contraindicated for surgical resection for multinodular HCC. Based on the tumor burden, the numerous subgroups [11; 12; 13; 14] had been identified to select a favorable treatment. The previous study showed that a higher LDH level was associated with worse outcomes after hepatectomy or TACE[22]. A correlation was also demonstrated between high serum LDH levels and high tumor volume, a high percentage of necrosis, or an aggressive phenotype for gastric and pancreatic cancer[23; 24]. In this study, we found a subgroup that HR was superior to TACE for intermediate-stage HCC in the range of LDH level > 192 U/L. Its underlying mechanism was still unclear, and one possible reason might be that surgery reduced the recurrence risk by removing larger lesions with a more aggressive phenotype.

Our study had some strengths. First, we created a propensity score-matched cohort to minimize potential bias. Next, our study provided a new insight for selecting the proper HCC patients to treat with surgical resection. The hematological indicators, such as LDH level, should be a promising biomarker.

Our study also had several limitations. Firstly, this was a retrospective cohort from real-world data; residual bias and unmeasured confounders were unavoidable, even if we had used the propensity score matching to eliminate inherent differences between two groups. Results after PS-matching and MI revealed that the bias from confounders and missing data might overestimate the advantage of surgical resection. On the contrary, this would make the benefit from TACE treatment more significant in the range of LDH level ≤ 192 U/L. Secondly, because this was a secondary analysis, the surgical program (radical vs. palliative, laparoscopic vs. open) was unclear. The difference in the cirrhosis rate, portal hypertension, and MELD score between the two groups was unknown, although the PS-matching results were consistent. Thirdly, this study focused on East Asia’s populations with hepatitis B between January 2007 and May 2012. Thus, our conclusions might not be suitable for western populations. With the development of more aggressive surgical treatment, the cut-off value of LDH should be further explored. In the future, the interaction between the effect of HR and LDH levels should be validated in the randomized control trial and larger scale real-world data in the various populations.

## 5. Conclusion

Hepatic resection was superior to TACE for intermediate-stage HCC in the range of LDH level > 192 U/L. Moreover, TACE might be suitable for patients with LDH level ≤ 192 U/L.

## Data Availability

the raw data were freely obtained from the Dryad Digital Repository database

https://doi.org/10.5061/dryad.pd44k8r

## Acknowledgments

Xiong Chen received financial support from the Natural Science Foundation of Fujian Province (Nos 2018J01352, 2016J01576, and 2016J01586); the Science and Technology Innovation Joint Foundation of Fujian Province(Nos 2017Y9125). We are grateful for the raw data from Prof. Peihong Wu and the statistical support from the Empower U team of the Department of Epidemiology and Biostatistics, X&Y solutions Inc. in Boston. Dr. Lu is grateful for his girlfriend’s kindness and support in the past four years, and Miss Lin, would you like to marry him?

