## supplemental materials for "Hepatic resection versus transarterial chemoembolization for the intermediate stage hepatocellular carcinoma: A cohort study" (from another version of this preprint: version 2, 2021-08-04)

**Supplementary materials**

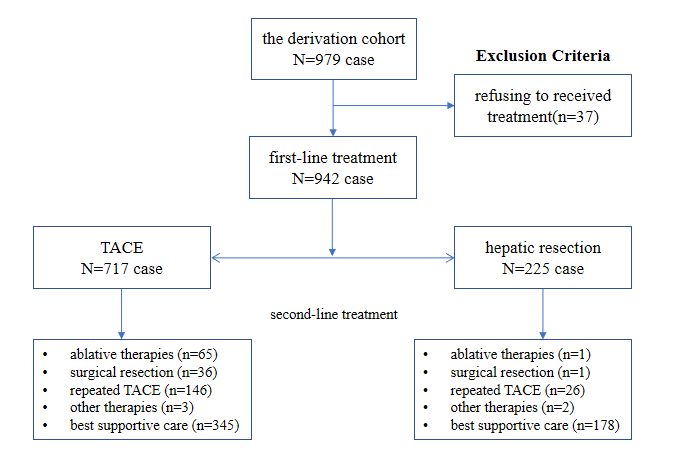

Fig S1. Flowchart of the derivation cohort.

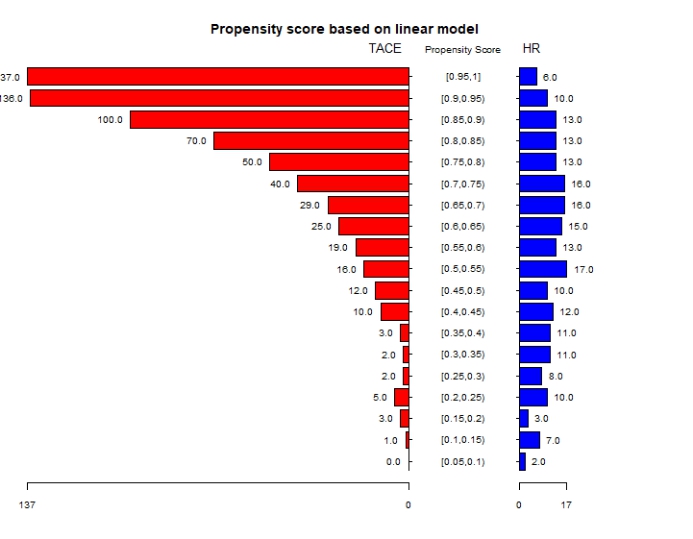

**Fig S2. Distribution of propensity-matching treatment strategy (TACE vs. HR) groups after matching.**

**Table S1. Baseline characteristics between TACE and HR group in the derivation cohort after propensity matching.**

|  | Treatment | | *P*-value |
| --- | --- | --- | --- |
|  | TACE | HR |  |
| No. | 169 | 169 |  |
| Age | 52.10 ± 12.72 | 51.57 ± 12.20 | 0.6946 |
| Gender |  |  | 0.1382 |
| male | 161 (95.3) | 153 (90.5) |  |
| female | 8 (4.7) | 16 (9.5) |  |
| HBV infection |  |  | 0.6504 |
| no | 4 (2.7) | 2 (1.3) |  |
| yes | 145 (97.3) | 152 (98.7) |  |
| Child-Pugh class |  |  | 0.6733 |
| A | 140 (82.8) | 136 (80.5) |  |
| B | 29 (17.2) | 33 (19.5) |  |
| Diameter of main tumor(cm) | 6.4 ± 3.3 | 6.4 ± 2.7 | 0.8308 |
| Both lobe with lesions |  |  | 0.2263 |
| no | 91 (53.8) | 103 (60.9) |  |
| yes | 78 (46.2) | 66 (39.1) |  |
| No. of intrahepatic lesions |  |  | 0.9121 |
| ≤3 | 100 (59.2) | 98 (58) |  |
| >3 | 69 (40.8) | 71 (42) |  |
| AFP(ng/ml) |  |  | 1 |
| <25 | 59 (34.9) | 59 (34.9) |  |
| ≥25 | 110 (65.1) | 110 (65.1) |  |
| CRP(mg/L) |  |  | 0.499 |
| <10 | 66 (39.1) | 59 (34.9) |  |
| ≥10 | 103 (60.9) | 110 (65.1) |  |
| Hgb(g/L) |  |  | 0.904 |
| <120 | 47 (27.8) | 49 (29) |  |
| ≥120 | 122 (72.2) | 120 (71) |  |
| LDH(U/L) |  |  | 0.9095 |
| <245 | 110 (65.1) | 108 (63.9) |  |
| ≥245 | 59 (34.9) | 61 (36.1) |  |
| WBC(10^9^/L) |  |  | 1 |
| <11 | 135 (79.9) | 135 (79.9) |  |
| ≥11 | 34 (20.1) | 34 (20.1) |  |
| PLT(10^9^/L) |  |  | 0.1534 |
| <150 | 103 (60.9) | 89 (52.7) |  |
| ≥150 | 66 (39.1) | 80 (47.3) |  |

**Table S2. Landmark analyses of overall survival for long-term (≥ 6 months, ≥ 1 year, ≥ 2 years) survivors.**

| Landmark | No. (%) | HR | 95%CI | *P*-value |
| --- | --- | --- | --- | --- |
| ≥6-month survivors | 204 (27.42%) | 0.45 | 0.35, 0.58 | <0.0001 |
| ≥1-year survivors | 174 (30.91%) | 0.46 | 0.34, 0.62 | <0.0001 |
| ≥2-year survivors | 127 (37.69%) | 0.52 | 0.33, 0.79 | 0.0027 |

**Table S3. Association between overall survival and platelet count /lactic dehydrogenase stratified by tertile for the intermediate-stage HCC.** (vs. TACE in the bottom tertile)

|  | Treatment | No. | HR | 95%CI | | *P*-value | P  for interaction |
| --- | --- | --- | --- | --- | --- | --- | --- |
|  |  |  |  | Low | High |  |  |
| **Before PS matching** |  |  |  |  |  |  |  |
| LDH (U/L) Tertile |  |  |  |  |  |  | 0.0062 |
| Low | TACE | 217 | 1.0 (ref.) | 1.0 (ref.) | 1.0 (ref.) |  |  |
| Low | HR | 95 | 0.65 | 0.45 | 0.93 | 0.0191 |  |
| Middle | TACE | 226 | 1.69 | 1.31 | 2.16 | <0.0001 |  |
| Middle | HR | 85 | 0.67 | 0.46 | 0.98 | 0.0395 |  |
| High | TACE | 267 | 2.02 | 1.59 | 2.57 | <0.0001 |  |
| High | HR | 45 | 0.46 | 0.25 | 0.84 | 0.0112 |  |
| PLT (10^9^/L) Tertile |  |  |  |  |  |  | 0.0371 |
| Low | TACE | 257 | 1.0 (ref.) | 1.0 (ref.) | 1.0 (ref.) |  |  |
| Low | HR | 54 | 0.54 | 0.34 | 0.85 | 0.0082 |  |
| Middle | TACE | 219 | 1.13 | 0.89 | 1.43 | 0.3022 |  |
| Middle | HR | 92 | 0.52 | 0.37 | 0.75 | 0.0004 |  |
| High | TACE | 233 | 1.75 | 1.4 | 2.2 | <0.0001 |  |
| High | HR | 79 | 0.47 | 0.31 | 0.69 | 0.0001 |  |
| **After PS matching** |  |  |  |  |  |  |  |
| LDH (U/L) Tertile |  |  |  |  |  |  | 0.0002 |
| Low | TACE | 60 | 1.0 (ref.) | 1.0 (ref.) | 1.0 (ref.) |  |  |
| Low | HR | 53 | 1.24 | 0.7 | 2.22 | 0.4613 |  |
| Middle | TACE | 54 | 2.76 | 1.61 | 4.71 | 0.0002 |  |
| Middle | HR | 58 | 1.4 | 0.79 | 2.48 | 0.251 |  |
| High | TACE | 55 | 3.72 | 2.17 | 6.4 | <0.0001 |  |
| High | HR | 58 | 0.84 | 0.45 | 1.58 | 0.5874 |  |
| PLT (10^9^/L) Tertile |  |  |  |  |  |  | 0.1195 |
| Low | TACE | 72 | 1.0 (ref.) | 1.0 (ref.) | 1.0 (ref.) |  |  |
| Low | HR | 40 | 0.69 | 0.39 | 1.24 | 0.2145 |  |
| Middle | TACE | 48 | 0.92 | 0.55 | 1.55 | 0.7637 |  |
| Middle | HR | 63 | 0.62 | 0.37 | 1.03 | 0.0637 |  |
| High | TACE | 49 | 1.88 | 1.16 | 3.06 | 0.0104 |  |
| High | HR | 66 | 0.63 | 0.38 | 1.04 | 0.0689 |  |

**Table S4. Association between HR and observed mortality at 1, 3, and 5 years stratified with LDH level.** (vs TACE)

| LDH (U/L) |  | OR(95%CI) | | |
| --- | --- | --- | --- | --- |
|  |  | 1 year | 3 year | 5 year |
| < 192 | 113 | 2.89 (0.71, 11.81) | 1.20 (0.54, 2.65) | 1.22 (0.57, 2.62) |
| ≥ 192 | 225 | 0.35 (0.18, 0.67) | 0.33 (0.19, 0.57) | 0.32 (0.18, 0.55) |

**Table S5. Association between overall survival and the confounding factors for the intermediate-stage HCC.**

|  | **Before PS matching** | | | **After PS matching** | | |
| --- | --- | --- | --- | --- | --- | --- |
|  | *No.* | HR(95%CI) | *P* for interaction | *No.* | HR(95%CI) | *P* for  interaction |
| Child-Pugh class |  |  | 0.737 |  |  |  |
| A | 799 | 0.42 (0.32, 0.54) |  |  |  |  |
| B | 143 | 0.38 (0.22, 0.65) |  |  |  |  |
| Diameter of main tumor(cm) |  |  | 0.713 |  |  |  |
| <5 | 300 | 0.35 (0.21, 0.57) |  |  |  |  |
| ≥5 | 642 | 0.43 (0.33, 0.56) |  |  |  |  |
| Lesions of lobe |  |  | 0.306 |  |  |  |
| Unilobar | 402 | 0.48 (0.35, 0.66) |  |  |  |  |
| Bilobar | 540 | 0.39 (0.27, 0.56) |  |  |  |  |
| No. of intrahepatic lesions |  |  | 0.362 |  |  |  |
| ≤3 | 379 | 0.48 (0.35, 0.66) |  |  |  |  |
| >3 | 563 | 0.41 (0.28, 0.58) |  |  |  |  |
| AFP(ng/ml) |  |  | 0.992 |  |  |  |
| <25 | 255 | 0.41 (0.27, 0.63) |  |  |  |  |
| ≥25 | 638 | 0.43 (0.33, 0.57) |  |  |  |  |
| LDH(U/L) Tertile |  |  | 0.006 |  |  | 0.0002 |
| 1 | 312 | 0.63 (0.44, 0.91) |  | 113 | 1.20 (0.67, 2.15) |  |
| 2 | 311 | 0.40 (0.27, 0.57) |  | 112 | 0.50 (0.30, 0.84) |  |
| 3 | 312 | 0.25 (0.14, 0.44) |  | 113 | 0.26 (0.14, 0.47) |  |
| PLT(10^9^/L) Tertile |  |  | 0.037 |  |  | 0.120 |
| 1 | 311 | 0.49 (0.35, 0.68) |  | 112 | 0.69 (0.39, 1.24) |  |
| 2 | 311 | 0.33 (0.24, 0.46) |  | 111 | 0.62 (0.35, 1.11) |  |
| 3 | 312 | 0.25 (0.14, 0.44) |  | 115 | 0.34 (0.20, 0.59) |  |
| CNLC stage |  |  | 0.362 |  |  |  |
| IIa | 379 | 0.47 (0.34, 0.65) |  |  |  |  |
| IIb | 563 | 0.41 (0.28, 0.58) |  |  |  |  |
